# PBMC transcriptional and plasma chemokine profiles show differential associations with the presence and burden of coronary artery calcification

**DOI:** 10.64898/2026.11.24.26368918

**Authors:** Andrea Domínguez-Igual, Laura Martín-Chaves, María Díaz-Ottaviano, Olga Popova, Lucía Beltrán-Camacho, María Lucía Berma-Gascón, Antonio Adarve-Castro, Rafael González-Manzanares, Germán Berteli-García, Francesco Costa, Ada del Mar Carmona-Segovia, Ana Isabel Molina-Ramos, Jorge Rodríguez-Capitán, Manuel F. Jiménez-Navarro, Francisco Javier Pavón-Morón

## Abstract

**Background:** Coronary artery calcium (CAC) reflects atherosclerotic burden. We examined whether peripheral blood mononuclear cell (PBMC) expression of calcium-regulatory and chemotactic-receptor genes and plasma chemokine concentrations were associated with CAC presence and with its extent among adults with detectable calcification.

**Methods:** This single-center, cross-sectional study included 53 adults undergoing coronary computed tomography, 27 with CAC=0 and 26 with CAC>0. We measured 10 PBMC transcripts by reverse transcription quantitative polymerase chain reaction and 8 plasma chemokines by multiplex immunoassay. We assessed associations using permutation tests, composite indices, and compartment-specific principal component analyses.

**Results:** All 10 PBMC transcripts correlated positively with CAC after false discovery rate correction. Associations were strongest for *S100A9* (rho=+0.69), *S100A8* and *CXCR2* (both rho=+0.66), and *CCR2* (rho=+0.65). Plasma CX_3_CL1, CXCL2, CXCL16, and CXCL12 were lower in participants with detectable CAC, whereas CXCL1 was higher and correlated with PBMC *CXCR2* expression (rho=+0.66, p<0.001). Most molecular differences distinguished participants with and without CAC. In exploratory analyses among participants with detectable CAC, *CCR2* expression (rho=+0.43, p=0.043), the chemotactic-receptor index (rho=0.51, p=0.022), and plasma CXCL1 (rho=+0.42, p=0.034) correlated with CAC burden. Additional adjustment for diabetes or hypertension attenuated selected associations.

**Conclusions:** Detectable coronary calcification was associated with coordinated differences in PBMC gene expression and a distinct plasma chemokine profile. *CCR2* expression, the receptor index, and plasma CXCL1 warrant further study in relation to the extent of calcification.

**What Are the Clinical Implications?:** In adults undergoing coronary computed tomography, the presence of coronary calcium was associated with differences in calcium-regulatory and chemotactic-receptor gene expression in circulating immune cells and in plasma chemokine concentrations. Among participants with detectable calcium, CCR2 expression, the chemotactic-receptor index, and plasma CXCL1 also tracked with calcium burden. These observations identify candidates for longitudinal studies testing whether circulating immune profiles are associated with subsequent changes in coronary calcium. Their value for individual risk assessment remains to be established.

## 1 Introduction

Coronary artery calcium (CAC) quantified by computed tomography is an established marker of subclinical atherosclerosis and cardiovascular risk and improves risk classification beyond traditional risk factors^1,2^. Vascular calcification is an active, regulated process involving inflammation, osteogenic differentiation of vascular cells, and the release of calcifying extracellular vesicles by macrophages^3–5^. Whether greater coronary calcification is associated with measurable differences in circulating immune-cell phenotypes remains incompletely defined.

Recruitment of circulating leukocytes to atherosclerotic lesions is regulated by several chemokine-receptor axes^6,7^. CCL2/CCR2 contributes to classical monocyte recruitment^8^, CX_3_CL1/CX_3_CR1 mediates nonclassical monocyte trafficking and endothelial adhesion^9^, CXCL1-CXCL2/CXCR2 participates in myeloid-cell mobilization and lesional macrophage accumulation^10^, and CXCL16/CXCR6 contributes to the positioning of lymphocytes and monocytes within atherosclerotic lesions^11^. Increased CX_3_CR1 expression has been reported in peripheral blood mononuclear cells (PBMCs) and monocytes from patients with coronary artery disease^12,13^, and a common CCR2 variant has been associated with the extent of coronary calcification in humans^14^.

Chemokine receptors can activate phospholipase C-β and IP3-dependent calcium release from endoplasmic reticulum stores^15^. Store depletion subsequently engages STIM1–ORAI1-mediated calcium entry, linking chemokine sensing to leukocyte migration, activation, and cytokine production^16,17^. Calreticulin (CALR) supports endoplasmic reticulum calcium buffering, and calmodulin (CALM1) transduces calcium-dependent signals^18^. S100A8 and S100A9 form calprotectin, are associated with myeloid activation, and have been implicated in mineral nucleation within macrophage-derived matrix vesicles^19,20^. These relationships provide a rationale for examining chemotactic and calcium-regulatory transcription together (**Figure S1**).

We hypothesized that coronary calcification would be associated with coordinated PBMC transcriptional differences involving calcium-regulatory and chemotactic pathways, accompanied by a distinct circulating chemokine profile. We examined these measures and their within- and cross-compartment relationships, distinguishing associations with CAC presence from variation in burden among CAC-positive participants.

## 2 Methods

Access to the data is restricted and available upon reasonable request. Researchers interested in accessing the dataset should contact the corresponding author.

### 2.1 Study design, participants, and clinical assessment

This was a single-center, cross-sectional observational study. Participants were consecutively recruited through the Department of Cardiology and Radiology at Hospital Universitario Virgen de la Victoria (Málaga, Spain) between January 2024 and December 2025. Eligible participants were adults aged ≥18 years who underwent coronary computed tomography for clinical indications and provided written informed consent. Exclusion criteria included active infectious or neoplastic disease, autoimmune disease or other chronic inflammatory conditions, severe cardiovascular disease, and cognitive or language limitations that could interfere with study participation.

Participants were classified as CAC-negative (score=0) or CAC-positive (score>0). CAC-positive participants were also grouped into observed CAC tertiles for descriptive visualization, exploratory comparisons, and trend tests. CAC remained continuous in the primary association analyses.

Medical records and interviews provided age, sex, body mass index, diabetes, dyslipidemia, hypertension, smoking status (never, current, or former), and prior cardiac events. These variables characterized the cohort and were used as covariates where specified.

The study was approved by the competent Research Ethics Committee through the Andalusian Biomedical Research Ethics Portal (PEIBA; protocol CAR06072022) and was conducted in accordance with the Declaration of Helsinki and applicable data-protection regulations. All participants provided written informed consent before study procedures. Clinical, sociodemographic, laboratory, and imaging data were coded using an alphanumeric identifier and stored in an anonymized research database.

### 2.2 Coronary calcium quantification

Noncontrast, prospectively ECG-triggered cardiac computed tomography used a multidetector scanner (Siemens Healthineers, Erlangen, Germany) at 120 kVp, with tube current adjusted to body habitus. Images were reconstructed at 2.5-mm slice thickness using a standard kernel and filtered back projection, at 75% of the R–R interval. Global CAC was quantified by the Agatston method and expressed in Agatston units (AU)^21^.

### 2.3 Sample collection and PBMC isolation

Fasting blood was collected in K2-EDTA tubes (BD Vacutainer, Becton Dickinson, Franklin Lakes, NJ, USA) on the computed tomography day and processed within 2 h. Density-gradient centrifugation with Ficoll separated plasma and PBMCs. Plasma aliquots were stored at −80 °C, and PBMCs were preserved for RNA extraction.

### 2.4 Gene expression by reverse transcription quantitative polymerase chain reaction (RT-qPCR)

PBMC RNA was extracted using TRIzol, treated with DNase, and reverse-transcribed. Ten transcripts (*CALR, ORAI1, STIM1, CALM1, S100A8, S100A9, CX3CR1, CCR2, CXCR2,* and *CXCR6*) were quantified in technical duplicate using SYBR Green RT-qPCR (Thermo Fisher Scientific, Waltham, MA, USA), following applicable MIQE recommendations^22^. Primer sequences and amplification characteristics are provided in **Tables S1** and **S2**. Additional procedures appear in the Supplemental Methods.

Expression was normalized to the arithmetic mean Ct of *ACTB* and *HPRT1*, equivalent to the geometric mean of their relative quantities^23^.ΔCt was target Ct minus mean reference Ct; results are reported as −ΔCt, with higher values indicating higher relative expression^24^.

Prespecified quality criteria excluded determinations when the mean reference-gene Ct exceeded 32 or a reference gene was undetermined. Eight participants failed these criteria (80 target determinations), and four additional target measurements were undetermined: 84/530 values (15.8%) were excluded, leaving 43–45 valid determinations per transcript. Low-abundance transcripts were interpreted cautiously (**Table S2**).

### 2.5 Plasma chemokines

Eight plasma chemokines (CX_3_CL1, CXCL1, CXCL2, CXCL10, CCL2, CCL7, CXCL16, and CXCL12) were measured in technical duplicate using Bio-Plex Pro Human Chemokine singleplex assays combined into an eight-analyte panel (Bio-Rad Laboratories, Hercules, CA, USA). Assay identifiers and detailed procedures are provided in the Supplemental Methods.

Plates were read on a Luminex 200 analyzer (Luminex, Austin, TX, USA), with personnel blinded to group assignment. Intra-assay and interassay coefficients of variation were <10% and <15%, respectively. Concentrations are reported in pg/mL.

### 2.6 Statistical analysis

Continuous variables are summarized as median with interquartile range (IQR), and categorical variables as n (%). Baseline continuous variables were compared using the Mann–Whitney U test, and categorical variables using Fisher’s exact test or the chi-square test, as appropriate.

Molecular analyses used two-sided permutation inference because groups were small and technical exclusions produced transcript-specific sample sizes^25^. Group comparisons, Spearman correlations, and ordered trends used 20,000 resamples; four-group comparisons used 10,000. Regression coefficients used Freedman–Lane residual permutation with 10,000 resamples^26^. Spearman confidence intervals used Fisher z transformation. Test statistics and permutation calculations are detailed in the Supplemental Methods.

Associations were examined in the full cohort and secondarily among CAC-positive participants. Benjamini–Hochberg false discovery rate (FDR) correction^27^ was applied separately to ten transcript correlations, eight chemokine correlations, and eight plasma group comparisons; adjusted probabilities are reported as q-values. Tertile trend and within-patient analyses were exploratory and unadjusted for multiple comparisons.

Three indices were calculated as mean standardized transcript scores in complete technically valid cases: Calcium signaling (*CALR, ORAI1, STIM1, CALM1, S100A8,* and *S100A9*), chemotactic receptors (*CX3CR1, CCR2, CXCR2*, and *CXCR6*), and *S100A8/A9*. Principal component analysis (PCA) was performed separately on standardized transcripts and log_2_-transformed, standardized chemokines. Plasma principal component 1 (PC1) was oriented toward higher CAC. PC1 scores were correlated in participants with complete data in both compartments; pairwise Spearman correlations were displayed as a matrix.

Linear models related each PBMC summary measure and log_2_-transformed chemokine concentration to log_2_(CAC+1), initially adjusting for age and sex. Diabetes and smoking were added separately. Exploratory plasma models considered hypertension alone or with diabetes, with FDR correction across eight hypertension-adjusted associations. Primary analyses used available cases; sensitivity analyses retained expression values without quality exclusions. Two-sided p<0.05 defined statistical significance. Analyses used Python 3.12 (NumPy, SciPy, statsmodels, and scikit-learn).

## 3 Results

### 3.1 Participant characteristics

A total of 53 adults were included, 27 controls with no detectable coronary calcium (CAC score=0) and 26 participants with detectable coronary calcification (CAC score >0). Among participants with detectable CAC, the observed score ranges across the three tertile groups were 66–374 AU (n=8), 503–974 AU (n=8), and 1237–5319 AU (n=10). Representative coronary computed tomography images are shown in **Figure S2A**.

Compared with controls, CAC-positive participants were older (66.5 vs 55.0 years), more often male (73% vs 37%), and had higher prevalences of diabetes (54% vs 7%), hypertension (81% vs 52%), and current or former smoking (58% vs 22%; **Table 1**). Dyslipidemia, prior cardiac events, and body mass index did not differ significantly. Median CAC in positive participants was 800.5 (334.0–1593.8) AU. The cohort and molecular analyses are summarized in **Figure S2B**, and CAC distributions in **Figure S2C**.

**Table 1.** Baseline characteristics of study participants.

| **Characteristic** | **Controls (n = 27)** | **Patients (n = 26)** | **p value** |
|---|---|---|---|
| Age, years (median and IQR) | 55.0 (51.5–59.5) | 66.5 (62.2–70.0) | < 0.001 |
| Male sex (n and %) | 10 (37) | 19 (73) | 0.013 |
| Body mass index, kg/m² (median and IQR) | 26.3 (24.3–28.6) | 29.2 (25.6–30.5) | 0.059 |
| Diabetes mellitus (n and %) | 2 (7) | 14 (54) | < 0.001 |
| Dyslipidemia (n and %) | 8 (30) | 15 (58) | 0.054 |
| Hypertension (n and %) | 14 (52) | 21 (81) | 0.042 |
| Smoking, current / former (n and %) | 5 (19) / 1 (4) | 3 (12) / 12 (46) | 0.002 |
| Prior cardiac event (n and %) | 0 (0) | 4 (15) | 0.051 |
| CAC score, AU (median and IQR) | 0 | 800.5 (334.0–1593.8) | — |

P values were obtained using the Mann–Whitney U test, Fisher's exact test, or chi-square test, as appropriate. AU, Agatston units; CAC, coronary artery calcium; IQR, interquartile range.

### 3.2 Calcium-regulatory transcripts and the CAC score

Median expression of all six calcium-regulatory transcripts was higher in participants with detectable CAC than in controls (**Table 2**; **Figure 1A**). Differences were largest for *S100A9* (10.09 vs 7.12 −ΔCt) and *S100A8* (1.40 vs −2.15; both p<0.001), followed by *CALR* (1.48 vs 0.35; p<0.001), *CALM1* (p=0.025), and *ORAI1* (p=0.011). *STIM1* showed the same direction of difference but did not reach statistical significance (p=0.055).

**Table 2.** PBMC gene expression and plasma chemokine concentrations by group and association with CAC score.

| **A. PBMC gene expression (−ΔCt)** |  |  |  |  |  |  |  |  |
| **Gene** | **Controls** | **Patients** | **Group p value** | **n** | **rho (95% CI)** | **p value** | **q value (FDR)** | **rho within patients** |
|---|---|---|---|---|---|---|---|---|
| *CALR* | 0.35 (0.03 to 0.89) | 1.48 (0.84 to 1.98) | < 0.001 | 45 | +0.57 (0.34 to 0.74) | < 0.001 | < 0.001 | +0.35 (p = 0.100) |
| *ORAI1* | −1.00 (−1.27 to −0.67) | −0.01 (−0.87 to 0.44) | 0.011 | 45 | +0.44 (0.17 to 0.65) | 0.002 | 0.003 | +0.38 (p = 0.076) |
| *STIM1* | −6.96 (−7.54 to −6.37) | −6.18 (−6.92 to −5.59) | 0.055 | 45 | +0.31 (0.02 to 0.55) | 0.041 | 0.041 | +0.16 (p = 0.458) |
| *CALM1* | −3.63 (−4.16 to −2.90) | −2.36 (−3.24 to −1.67) | 0.025 | 45 | +0.36 (0.07 to 0.59) | 0.016 | 0.018 | +0.22 (p = 0.322) |
| *S100A8* | −2.15 (−2.37 to −1.78) | 1.40 (−0.35 to 1.94) | < 0.001 | 45 | +0.66 (0.46 to 0.80) | < 0.001 | < 0.001 | +0.18 (p = 0.425) |
| *S100A9* | 7.12 (6.64 to 7.58) | 10.09 (9.52 to 11.54) | < 0.001 | 45 | +0.69 (0.50 to 0.82) | < 0.001 | < 0.001 | +0.28 (p = 0.190) |
| *CX3CR1* | −9.98 (−10.78 to −9.63) | −8.66 (−9.77 to −8.36) | 0.006 | 43 | +0.50 (0.23 to 0.69) | < 0.001 | 0.001 | +0.38 (p = 0.089) |
| *CCR2* | −5.08 (−5.51 to −4.33) | −2.78 (−3.85 to −1.27) | < 0.001 | 45 | +0.65 (0.44 to 0.79) | < 0.001 | < 0.001 | +0.43 (p = 0.043) |
| *CXCR2* | −9.54 (−10.09 to −8.78) | −7.23 (−8.30 to −7.05) | < 0.001 | 43 | +0.66 (0.44 to 0.80) | < 0.001 | < 0.001 | +0.34 (p = 0.139) |
| *CXCR6* | −10.51 (−11.40 to −10.02) | −9.70 (−10.60 to −8.93) | 0.023 | 45 | +0.38 (0.10 to 0.61) | 0.010 | 0.012 | +0.27 (p = 0.210) |
| **B. Plasma chemokine concentrations (pg/mL)** |  |  |  |  |  |  |  |  |
| **Chemokine** | **Controls** | **Patients** | **Group p value** | **rho (95% CI)** | **p value** | **q value (FDR)** | **Age + sex adjusted p value** |  |
| CX<sub>3</sub>CL1 | 134.21 (100.59 to 179.69) | 58.63 (35.02 to 77.15) | < 0.001 | −0.51 (−0.69 to −0.28) | < 0.001 | < 0.001 | 0.003 |  |
| CXCL1 | 31.15 (30.16 to 32.15) | 42.34 (39.11 to 43.83) | < 0.001 | 0.63 (0.43 to 0.77) | < 0.001 | < 0.001 | 0.024 |  |
| CXCL2 | 110.23 (38.75 to 170.38) | 24.43 (17.01 to 37.65) | < 0.001 | −0.44 (−0.63 to −0.19) | < 0.001 | 0.002 | 0.003 |  |
| CXCL10 | 153.00 (97.65 to 220.99) | 97.66 (58.67 to 206.34) | 0.063 | −0.24 (−0.48 to 0.03) | 0.084 | 0.111 | 0.589 |  |
| CCL2 | 36.82 (15.25 to 51.47) | 26.70 (20.69 to 39.07) | 0.370 | −0.09 (−0.36 to 0.18) | 0.501 | 0.501 | 0.591 |  |
| CCL7 | 275.49 (261.69 to 303.10) | 303.10 (265.14 to 339.35) | 0.192 | +0.22 (−0.06 to 0.47) | 0.119 | 0.136 | 0.635 |  |
| CXCL16 | 1360.9 (1021.9 to 1721.4) | 711.48 (642.80 to 979.72) | 0.001 | −0.44 (−0.63 to −0.19) | 0.001 | 0.002 | 0.036 |  |
| CXCL12 | 2767.4 (2491.3 to 3102.8) | 2380.6 (2119.9 to 2743.2) | 0.003 | −0.40 (−0.61 to −0.14) | 0.004 | 0.006 | 0.411 |  |

Values are median (IQR); expression is −ΔCt. n indicates valid determinations. Rho denotes full-cohort Spearman correlation with CAC; q values use Benjamini–Hochberg correction across ten genes. The final column reports correlations within CAC-positive participants. All p values use two-sided permutation tests (20,000 resamples). CAC, coronary artery calcium; CI, confidence interval; FDR, false discovery rate; IQR, interquartile range; PBMC, peripheral blood mononuclear cell.
Values are median (IQR), in pg/mL. Plasma was unavailable for one control. Rho denotes full-cohort Spearman correlation between log2 concentration and log2(CAC + 1); q values are corrected across eight chemokines. Group and correlation p values use 20,000 permutations; age- and sex-adjusted p values use Freedman–Lane permutation (10,000 resamples). CAC, coronary artery calcium; CI, confidence interval; FDR, false discovery rate; IQR, interquartile range.

**Figure 1.**
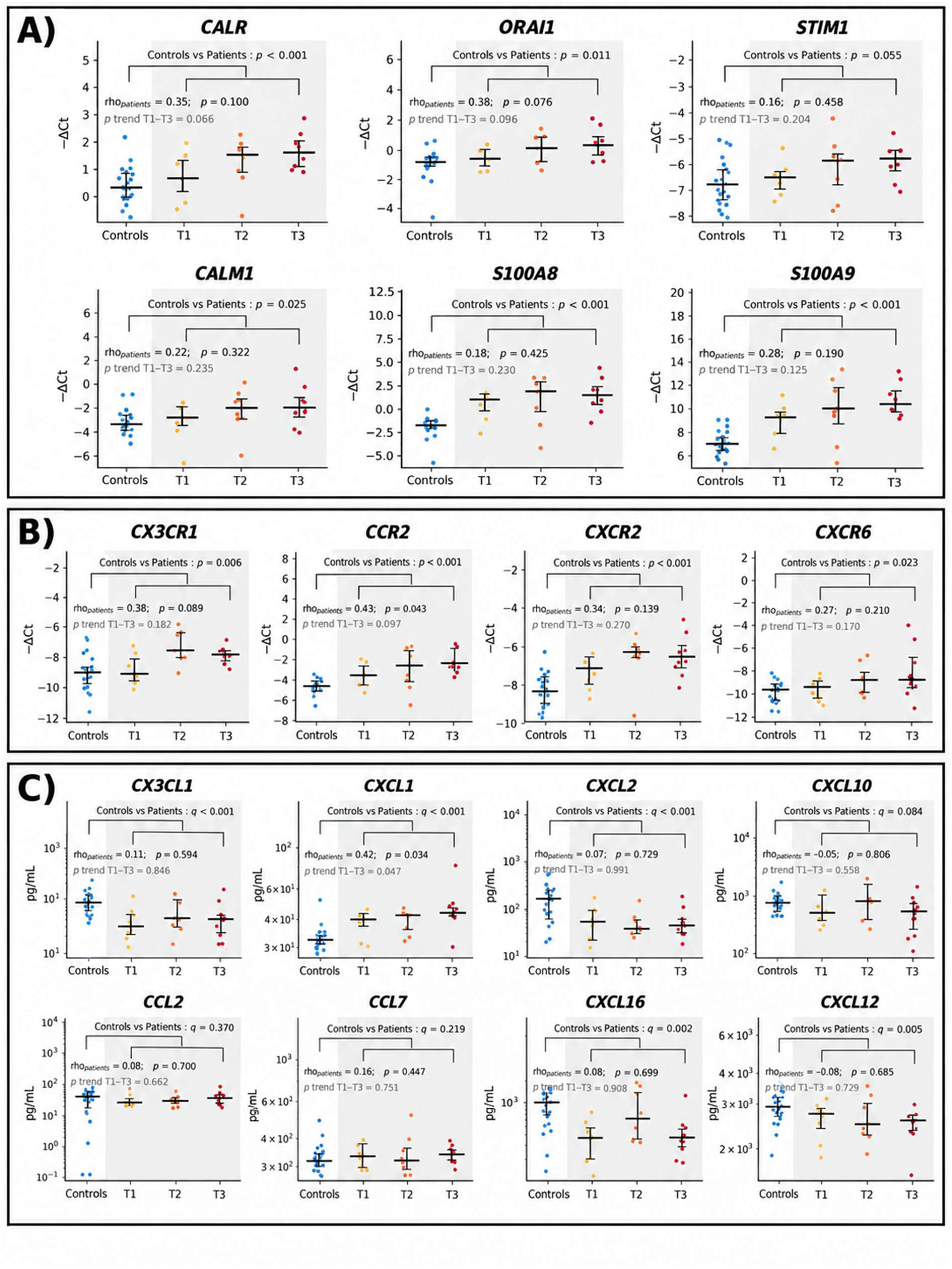
Expression of calcium-regulatory transcripts, chemotactic-receptor transcripts, and plasma chemokine concentrations across groups of coronary calcium burden. **(A)** Expression (−ΔCt) of *CALR*, *ORAI1*, *STIM1*, *CALM1*, *S100A8*, and *S100A9*; **(B)** Expression (−ΔCt) of *CX3CR1*, *CCR2*, *CXCR2*, and *CXCR6*; and (C) Concentrations (pg/mL; logarithmic scale) of CX_3_CL1, CXCL1, CXCL2, CXCL10, CCL2, CCL7, CXCL16, and CXCL12. Participants are shown as controls (CAC score=0) and as tertiles of the observed CAC distribution among participants with detectable calcification (T1, 66–374 AU; T2, 503–974 AU; T3, 1237–5319 AU). Symbols represent individual participants; horizontal bars indicate medians and vertical bars the IQR. Spearman correlations with continuous CAC score within participants with detectable calcification are shown in each panel. The p value for trend across CAC tertiles was obtained using the Jonckheere–Terpstra test and is presented descriptively. Group comparisons show nominal p values for PBMC transcripts and false discovery rate– adjusted q values for plasma chemokines. AU indicates Agatston units; CAC, coronary artery calcium; IQR, interquartile range; and PBMC, peripheral blood mononuclear cell.

In the full cohort, all six transcripts correlated positively with CAC score and remained significant after FDR correction. Correlations were strongest for *S100A9* (rho=+0.69) and *S100A8* (rho=+0.66), intermediate for *CALR* (rho=+0.57) and *ORAI1* (rho=+0.44), and weaker for *CALM1* (rho=+0.36) and *STIM1* (rho=+0.31). Expression differed across the four CAC groups for *CALR*, *S100A8*, and *S100A9* (all permutation Kruskal–Wallis p<0.001) and for ORAI1 (p=0.025), but not for *CALM1* (p=0.097) or *STIM1* (p=0.142) (**Table S3**).

### 3.3 Chemotactic-receptor transcripts and the CAC score

All four chemotactic-receptor transcripts were higher in participants with detectable CAC than in controls (**Table 2**; **Figure 1B**): *CCR2* (−2.78 vs −5.08 −ΔCt) and *CXCR2* (−7.23 vs −9.54; both p<0.001), *CX3CR1* (−8.66 vs −9.98; p=0.006), and *CXCR6* (−9.70 vs −10.51; p=0.023).

All four receptor transcripts correlated positively with CAC after FDR correction: *CXCR2* (rho=+0.66), *CCR2* (rho=+0.65), *CX3CR1* (rho=+0.50), and *CXCR6* (rho=+0.38). Four-group differences were significant for *CCR2*, *CXCR2*, and *CX3CR1*, but not *CXCR6* (**Table S3**). Overall, nine of ten transcripts differed between CAC-negative and CAC-positive participants; *STIM1* showed a similar but non-significant difference.

### 3.4 Plasma chemokines and the CAC score

Plasma chemokine concentrations differed between groups (**Table 2**; **Figure 1C**). CX_3_CL1 was lower in participants with detectable CAC than in controls (58.6 vs 134.2 pg/mL; p<0.001) and correlated inversely with CAC score (rho=−0.51; q<0.001); the association remained significant after adjustment for age and sex (p=0.003). CXCL2 (24.4 vs 110.2 pg/mL), CXCL16 (711.5 vs 1361.0 pg/mL), and CXCL12 (2380.6 vs 2767.4 pg/mL) were also lower in the CAC-positive group and correlated inversely with CAC (rho=−0.44, −0.44, and −0.40, respectively; all q≤0.006). CXCL2 (p=0.003) and CXCL16 (p=0.036) remained associated with CAC after adjustment for age and sex, whereas CXCL12 did not (p=0.411). CXCL1 was the only chemokine that was higher in participants with detectable CAC (42.3 vs 31.2 pg/mL; p<0.001). It correlated positively with CAC score (rho=+0.63; q<0.001) and remained associated after adjustment for age and sex (p=0.024). CXCL10, CCL2, and CCL7 did not differ significantly between groups. Among cognate ligand–receptor pairs, only CXCL1–*CXCR2* showed significant concordant cross-compartment covariation (rho=+0.66; p<0.001). Soluble CX_3_CL1 and PBMC *CX3CR1* expression were inversely correlated without reaching statistical significance (rho=−0.27; p=0.080), and no association was observed for CXCL2–*CXCR2* (rho=−0.14; p=0.382), CCL2–*CCR2* (rho=+0.04; p=0.793), or CXCL16–*CXCR6* (rho=−0.11; p=0.463) (**Figure 2**).

**Figure 2.**
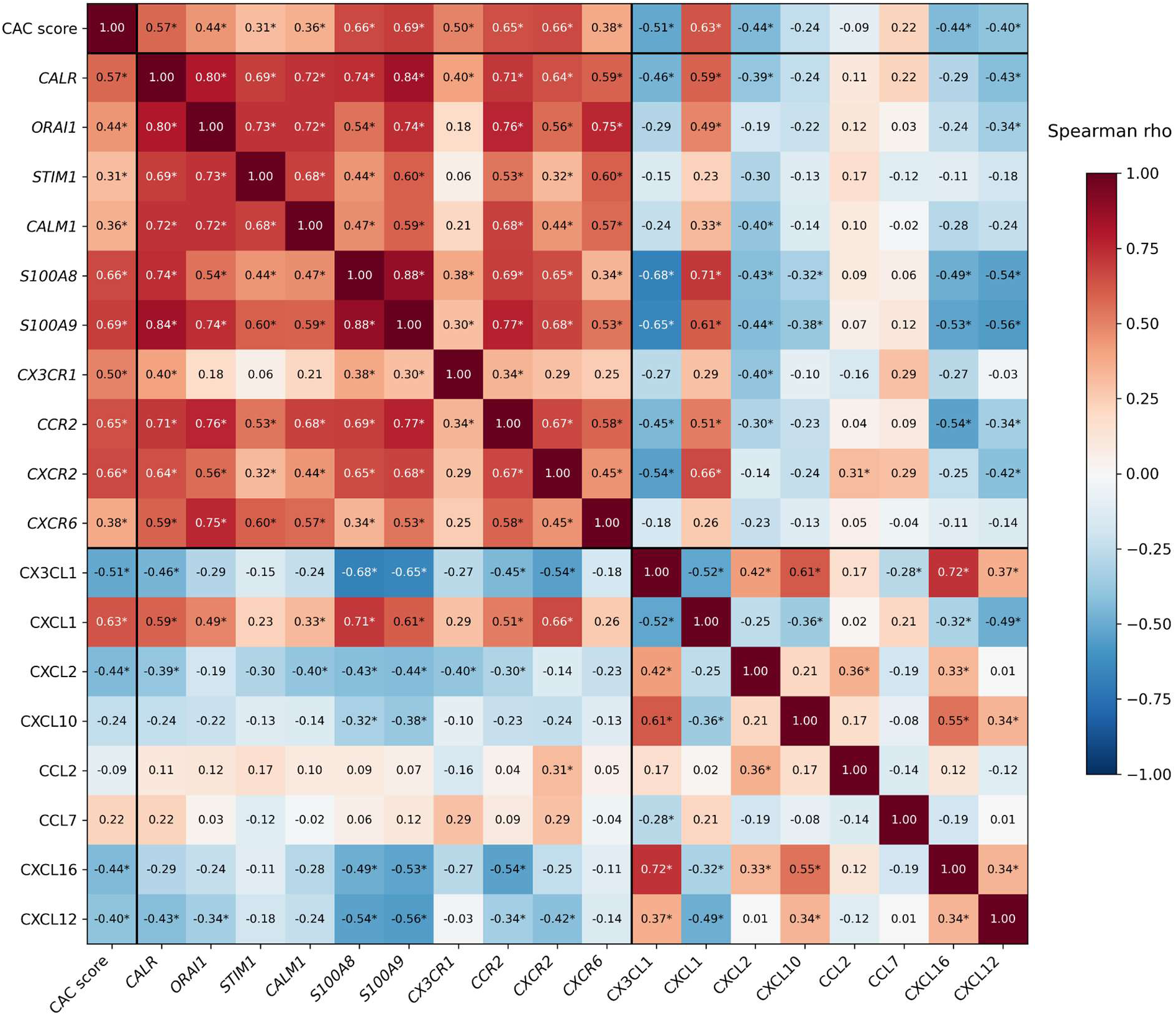
Spearman correlation matrix of CAC score, PBMC transcripts, and plasma chemokines. Pairwise Spearman correlations among CAC score, the ten PBMC transcripts (−ΔCt), and the eight plasma chemokines (log2-transformed) were computed using pairwise complete observations. Asterisks denote permutation p<0.05 without correction for multiple comparisons. Black lines separate CAC score, the PBMC compartment, and the plasma compartment. CAC indicates coronary artery calcium; and PBMC, peripheral blood mononuclear cell.

In sensitivity analyses additionally adjusted for hypertension, the associations of plasma CX_3_CL1, CXCL2, and CXCL16 with CAC remained nominally significant (p=0.025, 0.012, and 0.028, respectively), whereas that of CXCL1 was attenuated (p=0.109). None of these associations remained significant after FDR correction across the eight chemokines (all q≥0.074). With hypertension and diabetes included together, only CXCL2 retained a nominal association (p=0.035) (**Table S5**).

### 3.5 Association with the presence versus the burden of calcification

Ordered trends across patient tertiles and correlations restricted to CAC-positive participants distinguished group separation from variation within the positive CAC range (**Figure 1; Table S3**).

For most CAC-associated markers, the dominant difference was between CAC-negative and CAC-positive participants. *S100A8*, *S100A9*, and *CXCR2* separated the two groups clearly but varied little across patient tertiles, with no ordered trend (all p for trend across T1–T3 ≥0.12) and within-patient correlations of 0.18, 0.28, and 0.34, respectively (all p≥0.13). A similar pattern was evident in plasma: CX_3_CL1, CXCL2, CXCL16, and CXCL12 were already substantially shifted in the lowest CAC tertile and showed no monotonic decrease thereafter (all p for trend ≥0.72; within-patient rho between −0.08 and +0.11).

A smaller set of markers showed evidence of variation across the CAC-positive range. *CCR2* was the only individual transcript that remained significantly correlated with CAC among participants with calcification (rho=+0.43; p=0.043). Plasma CXCL1 was the only chemokine with a significant within-patient correlation (rho=+0.42; p=0.034) and the only chemokine with an ordered trend across tertiles (p for trend =0.047). *ORAI1* (rho=+0.38; p=0.076), *CX3CR1* (rho=+0.38; p=0.089), and *CALR* (rho=+0.35; p=0.100) showed positive but nonsignificant within-patient correlations, accompanied by similarly nonsignificant ordered trends across tertiles (p for trend =0.096, 0.182, and 0.066, respectively).

### 3.6 Integrated two-compartment profile

Correlations among the ten PBMC transcripts were uniformly positive, whereas correlations between PBMC transcripts and plasma chemokines were predominantly negative, with CXCL1 as the main exception (**Figure 2**). Among the 42 participants with complete technically valid transcript data, PBMC PC1 explained 58% of total variance and PC2 a further 13%. All ten transcripts loaded positively on PC1 (0.17–0.37) (**Figure 3A,B**).

**Figure 3.**
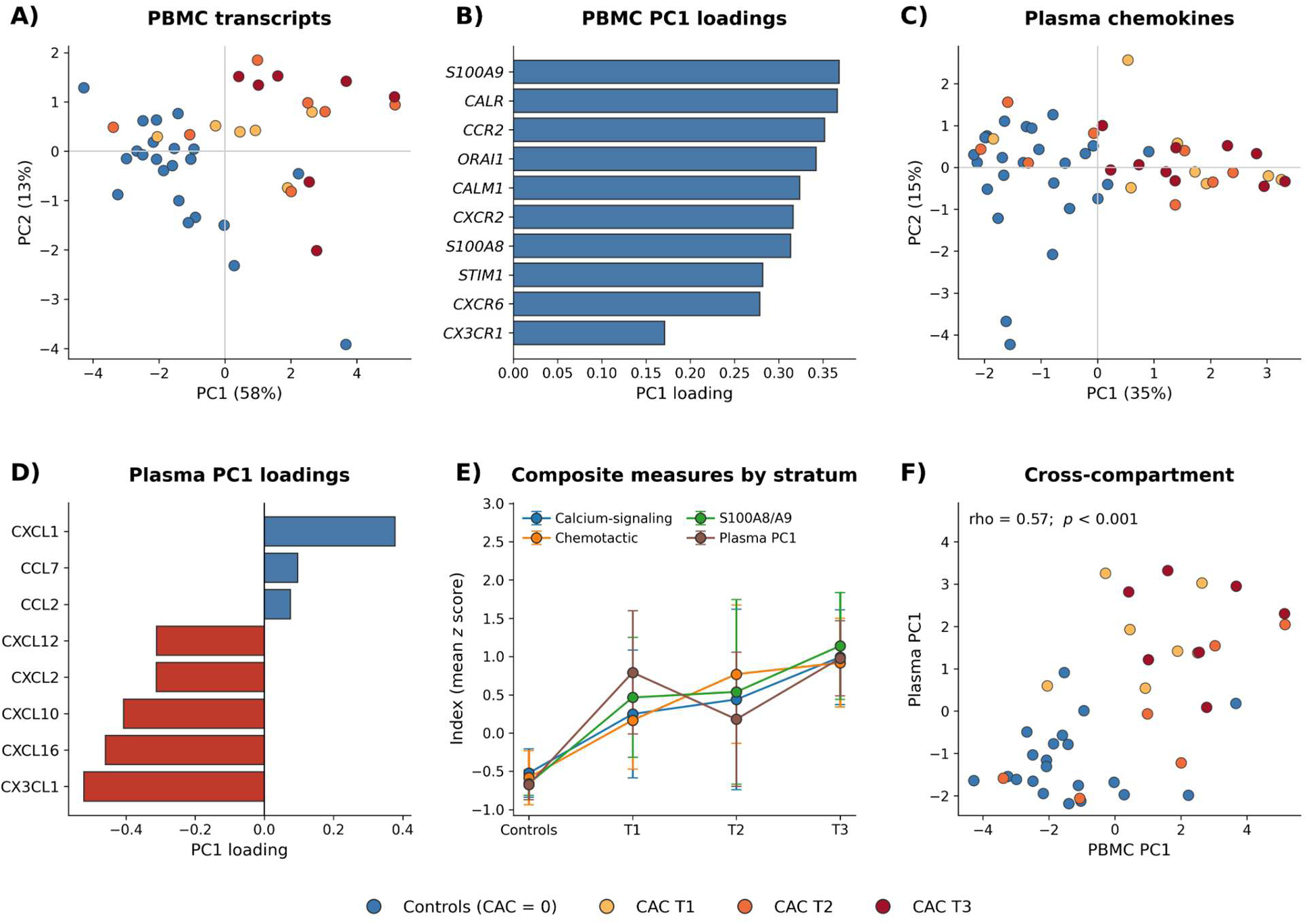
Two-compartment integrated profile. **(A)** PCA of the ten PBMC transcripts in participants with complete technically valid data, coded by CAC group. **(B)** PC1 loadings for the ten transcripts. **(C)** PCA of the eight log2-transformed plasma chemokines. **(D)** Plasma PC1 loadings, oriented so that higher PC1 values correspond to higher CAC burden. **(E)** Calcium-signaling, chemotactic-receptor, and *S100A8/A9* indices, together with standardized plasma PC1, across controls and the three observed CAC tertiles; symbols indicate means with 95% CI. **(F)** PBMC PC1 plotted against plasma PC1 in participants with complete data for both compartments. CAC, coronary artery calcium; PCA, principal component analysis; PC, principal component. CI indicates confidence interval; and PBMC, peripheral blood mononuclear cell.

PCA of the eight plasma chemokines showed a different pattern (**Figure 3C,D**). PC1 accounted for 35% of the total variance and PC2 for a further 15%. Plasma PC1 was characterized by opposing loadings for CX_3_CL1, CXCL16, CXCL10, CXCL2, and CXCL12 (negative) and CXCL1 (positive), whereas CCL2 and CCL7 contributed relatively little. PC2 was mainly associated with CCL2 and CCL7 and was unrelated to CAC score (rho=+0.07; p=0.610).

The calcium-signaling, chemotactic-receptor, and *S100A8/A9* indices, together with PBMC PC1, were higher in participants with detectable CAC (all p<0.001), correlated with CAC score (rho=+0.61, +0.71, +0.67, and +0.64, respectively; all p<0.001), and differed across the four CAC groups (permutation Kruskal–Wallis p<0.001) (**Table 3**; **Figure 3E**). The calcium-signaling and chemotactic-receptor indices were strongly correlated (rho=+0.79). Plasma PC1 was also higher in participants with detectable CAC (p<0.001), correlated with CAC score in the full cohort (rho=+0.64; p<0.001), and differed across CAC groups (p<0.001). However, plasma PC1 was not associated with CAC score among CAC-positive participants (rho=+0.03; p=0.870) and showed no ordered trend across tertiles (p for trend=0.796).

**Table 3.** Composite PBMC expression indices and coronary calcium burden.

| **Index** | **Controls** | **Patients** | **p value** | **rho (cohort)** | **rho (patients)** | **β (95% CI), age + sex** | **p value** | **β (95% CI), age + sex + diabetes** | **p value** |
|---|---|---|---|---|---|---|---|---|---|
| Calcium-signaling index | -0.51 (-0.84 to -0.24) | 0.66 (0.13 to 0.96) | < 0.001 | +0.61 (p < 0.001) | +0.32 (p = 0.167) | 0.11 (0.04 to 0.18) | 0.003 | 0.09 (0.01 to 0.18) | 0.030 |
| Chemotactic-receptor index | -0.56 (-0.81 to -0.23) | 0.44 (0.12 to 0.75) | < 0.001 | +0.71 (p < 0.001) | +0.51 (p = 0.022) | 0.07 (0.02 to 0.13) | 0.015 | 0.06 (-0.01 to 0.14) | 0.073 |
| *S100A8/A9* index | -0.63 (-0.80 to -0.49) | 0.79 (0.36 to 1.14) | < 0.001 | +0.67 (p < 0.001) | +0.22 (p = 0.347) | 0.12 (0.05 to 0.19) | 0.001 | 0.09 (0.00 to 0.17) | 0.042 |
| PC 1 | -1.56 (-2.40 to -0.96) | 1.75 (0.45 to 2.68) | < 0.001 | +0.64 (p < 0.001) | +0.40 (p = 0.081) | 0.31 (0.11 to 0.51) | 0.004 | 0.25 (0.01 to 0.49) | 0.039 |

Index values are median with IQR of the mean z score of the component genes; PC1 scores are reported in principal-component units. All p values are two-sided permutation p values (20,000 resamples for group comparisons and correlations; 10,000 Freedman–Lane resamples for regression coefficients). Rho, Spearman correlation with the CAC score in the full cohort and among participants with detectable CAC and valid data; β, change in the index per doubling of (CAC + 1), modelled as log<sub>2</sub>(CAC + 1), after adjustment for the indicated covariates. Permutation Kruskal–Wallis tests across the four CAC groups were significant for all four measures (p < 0.001). CAC, coronary artery calcium; IQR, interquartile range; PBMC, peripheral blood mononuclear cells; PC, principal component.

PBMC and plasma PC1 scores correlated in participants with complete data in both compartments (rho=+0.57; p<0.001; **Figure 3F**). Plasma CXCL1 correlated positively with *S100A8*, *CXCR2*, and *S100A9* (rho=+0.71, +0.66, and +0.61), whereas CX_3_CL1 correlated inversely with the same transcripts (rho=−0.68, −0.54, and −0.65). CCL2 and CCL7 showed no consistent associations with PBMC transcripts or CAC (**Figure 2**).

All four PBMC summary measures remained associated with CAC after adjustment for age and sex (**Table 3**). Adding diabetes attenuated these associations: the calcium-signaling index, *S100A8/A9* index, and PBMC PC1 remained significant (p=0.030, 0.042, and 0.039), whereas the receptor index did not (p=0.073). Adding smoking instead did not materially change the estimates. Among CAC-positive participants, the receptor index correlated with CAC (rho=+0.51; p=0.022); PBMC PC1 did not reach significance (rho=+0.40; p=0.081), nor did the calcium-signaling or *S*1*00A8/A9* indices (p=0.167 and 0.347).

### 3.7 Sensitivity analyses

Because the eight samples excluded by the quality-control criteria showed measurable rather than absent amplification, the principal sensitivity analysis retained all available determinations regardless of reference-gene Ct (n=44–52 per transcript). Under this less restrictive approach, seven of ten transcripts remained significantly correlated with CAC after FDR correction, whereas *CALM1*, *STIM1*, and *CXCR6* did not (**Table S4**). Effect estimates were generally attenuated (for example, *S100A9* rho=+0.60 vs +0.69 and *CALR* rho=+0.49 vs +0.57 in the primary analysis), but the direction of the associations was unchanged.

## 4 Discussion

In this cross-sectional study, coronary artery calcification was associated with a coordinated PBMC transcriptional profile involving calcium-regulatory proteins and chemotactic receptors, accompanied by a distinct pattern of circulating chemokines. The strongest transcript-level associations with CAC involved *S100A9*, *S100A8*, *CXCR2*, *CCR2*, and *CALR*. These transcripts showed substantial covariance, and the calcium-signaling and chemotactic-receptor indices were strongly correlated. In plasma, CX_3_CL1, CXCL2, CXCL16, and CXCL12 were lower in participants with detectable CAC, whereas CXCL1 was higher. Although most molecular differences primarily distinguished participants with and without detectable calcification, *CCR2*, the chemotactic-receptor index, and plasma CXCL1 also showed associations with CAC burden among CAC-positive participants. Sensitivity analyses showed attenuation of selected associations after accounting for diabetes or hypertension.

Among the PBMC transcripts, *S100A8* and *S100A9* showed particularly strong associations with CAC and favorable amplification characteristics. These genes encode the calcium-binding proteins that form calprotectin, a complex abundant in activated myeloid cells and involved in inflammatory signaling through TLR4 and RAGE^19,28^. Circulating calprotectin has also been associated with cardiovascular outcomes following acute coronary syndromes^29,30^. Experimental studies provide a further connection with vascular mineralization: S100A9 interacts with annexin V and phosphatidylserine in macrophage-derived matrix vesicles, facilitating hydroxyapatite nucleation, whereas S100A9 deficiency reduces their calcification potential^20^. In our cohort, however, *S100A8* and *S100A9* expression differed markedly according to the presence of CAC but showed little variation across the CAC-positive range. These findings are compatible with a broader myeloid inflammatory phenotype accompanying coronary atherosclerosis rather than a transcriptional response proportional to calcium burden. Because PBMC isolation largely excludes granulocytes, monocytes are a plausible source of these transcripts, although their cellular origin cannot be established without cell-specific analyses.

The calcium-regulatory transcripts also showed positive associations with CAC, particularly *CALR* and *ORAI1*, while *STIM1* and *CALM1* exhibited weaker associations. *CALR* contributes to endoplasmic reticulum calcium storage and protein homeostasis, whereas *STIM1* and *ORAI1* participate in store-operated calcium entry, a mechanism involved in immune-cell activation, migration, and cytokine production^16–18^. The positive loadings of all ten transcripts on PBMC PC1 and the strong correlation between the calcium-signaling and chemotactic-receptor indices suggest substantial coordination across these transcriptional domains. Such covariance is biologically plausible because chemokine receptors can engage PLCβ-dependent calcium mobilization and downstream calcium-sensitive signaling pathways^15–17^. Nevertheless, the calcium-signaling index also includes *S100A8* and *S100A9*, which are closely related to myeloid activation. Thus, its association with CAC may reflect overlapping calcium-regulatory and inflammatory transcriptional processes rather than a specific alteration in intracellular calcium homeostasis.

Among the chemotactic receptors, *CCR2* and *CXCR2* showed the strongest associations with CAC in the full cohort. Notably, *CCR2* was the only individual PBMC transcript that remained significantly correlated with CAC among participants with detectable calcification. CCR2 contributes to inflammatory monocyte recruitment during atherogenesis^8,31,32^, and a CCR2 polymorphism has previously been associated with coronary calcium burden^14^. The chemotactic-receptor index also retained an association within the CAC-positive subgroup, suggesting that variation in this transcriptional domain may extend beyond the distinction between calcified and noncalcified coronary arteries. Given the limited sample size and exploratory nature of these subgroup analyses, however, these associations require independent confirmation.

Plasma CXCL1 showed a different pattern from the other circulating chemokines. It was the only measured chemokine with higher concentrations in CAC-positive participants and a significant correlation with CAC within this subgroup. CXCL1 also correlated positively with PBMC *CXCR2* expression, representing the only cognate ligand– receptor pair with significant cross-compartment covariation in this study. This relationship is of particular interest because CXCR2 signaling can influence intracellular calcium mobilization and inflammatory-cell recruitment^10,15^. However, *CXCR2* expression itself was not significantly associated with CAC burden within the CAC-positive subgroup. The observed CXCL1–*CXCR2* relationship therefore supports coordinated variation between plasma and PBMC measurements but does not establish receptor activation or a common relationship with calcification severity.

The lower plasma concentrations of CX_3_CL1 and CXCL16 in CAC-positive participants warrant consideration because both chemokines have membrane-associated and soluble forms. CX_3_CL1 can mediate leukocyte adhesion through its membrane-bound form and is released by ADAM10- and ADAM17-dependent cleavage^33–35^. Previous studies have reported increased CX_3_CL1 expression or circulating concentrations in coronary artery disease, particularly in clinically unstable settings^13^. The lower soluble CX_3_CL1 concentrations observed here may reflect differences in vascular expression, proteolytic release, or tissue distribution, although these mechanisms cannot be distinguished using plasma measurements alone. CXCL16, which is also released through proteolytic shedding^36^, showed a similar reduction and remained associated with CAC after adjustment for age and sex. However, CXCL2 and CXCL12, which are not membrane-anchored chemokines, were also lower in CAC-positive participants, suggesting that altered shedding cannot account for the entire plasma profile. The inverse association of CXCL12 with CAC differs in direction from genetic evidence linking higher circulating CXCL12 to coronary artery disease^37^. Differences between circulating concentrations, local vascular signaling, and the clinical phenotypes examined may contribute to this discrepancy. In particular, CXCL12 distribution is influenced by endothelial binding and scavenging through ACKR3/CXCR7^38^, processes that were not assessed in this study.

The integrated analyses further distinguished the PBMC and plasma compartments. PBMC PC1 captured a substantial proportion of shared transcriptional variation, whereas plasma PC1 represented opposing contributions from CXCL1 and several other chemokines. Although both components were associated with CAC in the full cohort, their behavior differed within the CAC-positive subgroup. Plasma PC1 showed no association with CAC burden, whereas the chemotactic-receptor index retained a positive correlation. PBMC and plasma PC1 scores were themselves correlated, consistent with shared variation across the two molecular compartments. However, this relationship may partly reflect the differences between CAC-negative and CAC-positive participants and does not demonstrate direct functional coupling. Together, these findings suggest that circulating chemokine concentrations and PBMC transcriptional measures capture related but nonidentical aspects of the peripheral immune profile associated with coronary calcification.

### Strengths and limitations

Strengths include joint assessment of PBMC transcripts, plasma chemokines, and computed tomography-derived CAC, with analyses distinguishing CAC presence from burden. Prespecified amplification quality criteria, correction for multiple testing, permutation inference, and sensitivity analyses supported assessment of the associations.

Several limitations should nevertheless be acknowledged. The cross-sectional design does not establish the direction or causal nature of the relationships, and the modest sample size limited statistical power, particularly for analyses restricted to participants with detectable CAC. These subgroup analyses were exploratory and were not corrected for multiple comparisons. Participants with and without CAC also differed in age, sex, diabetes, hypertension, and smoking history. Age and sex were included in the primary adjusted models, with diabetes, smoking status, and hypertension considered in additional sensitivity analyses as applicable. Several associations were sensitive to these adjustments, although the direction of the main plasma associations was generally maintained. Residual confounding and instability of models containing several correlated cardiovascular risk factors remain possible.

Technical considerations also warrant attention. Low-abundance transcripts amplified near the assay’s limit of quantification, and measurements failing the predefined quality criteria were excluded from the primary analyses. Sensitivity analyses including all available determinations yielded associations in the same direction but generally smaller effect estimates. Although expression was normalized using reference genes, their stability across study groups was not independently validated. Furthermore, gene expression was measured in total PBMCs, precluding discrimination between changes in transcription within individual cell populations and differences in leukocyte subset composition^39^ . The correlation between PBMC and plasma principal components may also be influenced by group differences rather than reflecting an independent cross-compartment relationship. Finally, neither receptor activity nor intracellular calcium signaling was directly measured, and the absence of non-calcified plaque characterization prevents determining whether these molecular differences are specific to calcification or reflect broader coronary atherosclerotic burden.

### Conclusions

Coronary artery calcification was associated with coordinated PBMC transcriptional changes involving calcium-regulatory and chemotactic-receptor genes, together with a distinct circulating chemokine profile. Most molecular differences were associated with the presence of detectable calcification, whereas *CCR2*, the chemotactic-receptor index, and plasma CXCL1 also showed exploratory associations with CAC burden among affected participants. The covariance between transcriptional domains and the CXCL1–*CXCR2* cross-compartment relationship identify molecular associations that warrant further investigation using larger cohorts and cell-specific functional analyses.

## Acknowledgments

The authors thank Carolina Lobo from the Proteomics Unit at the Central Research Support Services of the University of Malaga for her technical support in the plasma samples processing and evaluation, and Ana María Pérez Sánchez for her contribution to the RT-qPCR analysis of PBMC gene expression carried out as part of her Bachelor’s thesis at the University of Málaga.

The authors used ChatGPT (OpenAI) for language editing and assistance with manuscript formatting and the wording of journal-specific sections. The authors are responsible for the content and references.

## Author Contributions

A.D.-I., A.d.M.C.-S., A.I.M.-R. contributed to Data curation; L.M.-C., M.D.-O., O.P., M.L.B.-G., A.A.-C., G.B.-G., and A.d.M.C.-S. contributed to Investigation and Methodology; L.M.-C., M.D.-O., O.P., L.B.-C., R.G.-M., and F.J.P.-M. contributed to Formal analysis; A.D.-I., F.C., J.R.-C., and A.I.M.-R. contributed to Validation; L.M.-C. and F.J.P.-M. contributed to Visualization; A.I.M.-R., M.F.J.-N., and F.J.P.-M. contributed to Conceptualization and Supervision; M.F.J.-N. and F.J.P.-M. contributed to Project administration; All authors contributed to Writing-original draft.

## Supplemental Material

Supplemental Methods

Figures S1–S2

Tables S1–S5

## Non-standard Abbreviations and Acronyms

AU: Agatston units
CAC: coronary artery calcium
Ct: quantification cycle
FDR: false discovery rate
MIQE: Minimum Information for Publication of Quantitative Real-Time PCR Experiments
PBMC: peripheral blood mononuclear cell
PC1: first principal component
PC2: second principal component
PCA: principal component analysis
RT-qPCR: reverse transcription quantitative polymerase chain reaction

